# Clinical characteristics and COVID-19 outcomes in a regional cohort of pediatric patients with rheumatic diseases

**DOI:** 10.1101/2021.05.19.21257450

**Authors:** Daniel Clemente, Clara Udaondo, Jaime de Inocencio, Juan Carlos Nieto, Pilar Galán del Río, Antía García Fernández, Jaime Arroyo Palomo, Javier Bachiller-Corral, Juan Carlos Lopez Robledillo, Leticia Leon, Lydia Abasolo, Alina Boteanu

**Author notes:** AB and LA shared senior authorship. **Corresponding author**: Leticia Leon, Rheumatology, IdISCC, Hospital Clínico San Carlos; C/ Martin Lagos s/n, 28040, Madrid, Spain. /3414.

## Abstract

**Background:** This study aimed to assess the baseline characteristics and clinical outcomes of coronavirus disease 2019 (COVID-19) in pediatric patients with rheumatic and musculoskeletal diseases (RMD) and identify the risk factors associated with symptomatic or severe disease defined as hospital admission, intensive care admission or death.

**Methods:** An observational longitudinal study was conducted during the first year of pandemic SARS-CoV-2 (1^st^ March 2020 to 1^st^ March 2021). All pediatric patients attended at the rheumatology outpatient clinic of six tertiary hospital in Madrid, Spain, with a medical diagnosis of RMD and COVID-19 were included. Main outcomes were symptomatic disease and hospital admission. The covariates were sociodemographic, clinical, and treatments. We ran a multivariable logistic regression model to assess risk factors for outcomes.

**Results:** The study population included 77 pediatric patients. Mean age was 11.88 (4.04) years Of these, 30 patients were asymptomatic, 41 had a mild or moderate disease and other 6 patients (7.79%) required hospital admission related to COVID-19. The median length of stay was 5 (2–20) days and there was no death. Previous comorbidities increased the risk for symptomatic disease and hospital admission. Compared with outpatients, the factor independently associated with hospital admission was the use of glucocorticoids (OR 1.08; p=0.00). No statistically significant findings for symptomatic COVID-19 were found in the final model.

**Conclusion:** Our data found no differences in COVID-19 outcomes between children-onset rheumatic diseases. Our results suggest that associated comorbidities and being in treatment with glucocorticoids increase the risk of hospital admission.

## Background

Severe acute respiratory syndrome coronavirus 2 (SARS-CoV-2) causes a myriad of clinical signs and symptoms, together with typical laboratory abnormalities, that manifest as the disease COVID-19 (1).

Since the confirmation of the first patient infected with SARS-CoV-2 in Spain in January 2020, the current COVID-19 outbreak has had a considerable impact, especially in the Madrid region, where a higher incidence of COVID-19 cases has been recorded (2).

The incidence and severity of COVID-19 disease seems to be higher in patients with risk factors, such as advanced age and associated comorbidities, mainly hypertension, diabetes, heart disease, and previous respiratory diseases (3)(4)(5). In this sense, having an autoimmune systemic condition could be considered another risk factor for severity since these patients had a higher risk of hospital admission related to COVID-19 (6).

In contrast with infected adults, most infected children appear to have a milder clinical course, and asymptomatic infections are not uncommon(7). Indeed, the admission rate to intensive care unit is 2–3% in pediatric patients, and fatal cases are rare (0,08%)(8)(9).

It is known that children can specific develop a multisystemic inflammatory syndrome during the COVID-19 clinical course (10). However, information of a more severe clinical course in patients with autoimmune or autoinflammatory condition is scarce.

Regarding therapies, studies in adult patients with rheumatic and musculoskeletal diseases (RMD) show that most of synthetic and biological disease-modifying antirheumatic drugs is not associated with a worse evolution of COVID-19. However, when patients have a poorly controlled active RMD or receive some treatments such as glucocorticoids, they may have an increased risk of both infection and a severe disease [(6)(11)].

There is a growing evidence that pediatric patients with autoimmune/inflammatory diseases receiving immunomodulatory therapy are not substantially endangered by SARS-CoV-2 beyond the common risk of viral infections (12). Besides, there is no reliable evidence to suggest a more severe disease in these patients.

Prior to this report, there had been several small case series of COVID-19 in pediatric patients with RMD reported (13). Evidence on the risk factors of poor outcome with COVID-19 specific to pediatric rheumatic disease is scarce. In addition, there are few data on how the hospital admissions of these patients with severe COVID-19 infection have evolved.

Therefore, due to a paucity of research data about COVID-19 in pediatric patients with rheumatic diseases, we conducted this study to obtain a general picture of pediatric patients with RMDs and COVID-19 in the region of Madrid. Thus, the purpose was to assess the baseline characteristics and clinical outcomes of COVID-19 in these patients and determine the risk factors associated with the development of mild, moderate or severe COVID-19.

## Methods

The COVID-SORCOM register is a multicenter regional register aiming to describe the clinical characteristics and outcomes of COVID-19 in patients with RMD in the Province of Madrid, Spain. We performed a retrospective observational study from the databases of the children and adolescents with RMD included in this register.

During the period between March 1, 2020, and March 1, 2021, clinical and demographic characteristics, COVID-19 data, and outcomes of these patients were retrospectively collected.

The inclusion criteria were age <18 years, a medical diagnosis (according to ICD-10) of a pediatric RMD, and COVID-19 disease confirmed with a positive SARS-CoV-2 polymerase chain reaction (PCR) test, positive rapid antigen test or detection of antibodies against SARS-CoV-2, or clinical diagnosis based on typical symptoms and epidemiological data during the first weeks of the pandemic, due to the lack of widely available diagnostic tests.

The severity of COVID-19 was defined on the basis of the clinical features, laboratory testing, and chest radiograph imaging, distinguishing between asymptomatic infection and symptomatic infection, further classified as mild, moderate, severe, or critical (14). We consider patients with mild disease those who had symptomatic infection but did not need admission; patients with moderate disease to those who required hospital admission due to symptoms, patients with severe disease if, in addition to requiring hospital admission, they needed an intensive care unit, and patients with critical disease those who had a poor outcome with significant sequelae or death.

Primary outcomes of interest were the symptomatic COVID-19 development and hospital admission due to COVID-19.

The co-variables recorded were as follows: **1)** Sociodemographic baseline characteristics including sex, age, and RMD duration. **2)** Type of RMD, classified into the following 3 groups: a) chronic inflammatory arthritis: juvenile idiopathic arthritis (JIA) (excluding the systemic subtype), enthesitis-related arthritis, and anterior chronic uveitis associated to JIA; b) multisystemic autoimmune disorders: systemic lupus erythematosus, idiopathic inflammatory myopathy, scleroderma and pediatric vasculitis and c) autoinflammatory diseases: systemic JIA, PFAPA syndrome, chronic recurrent multifocal osteomyelitis and monogenic autoinflammatory syndromes (TRAPS, CAPS, FMF, MKD). **3)** Disease activity according to the criteria of the responsible physician (clinical remission on and off medication, low, moderate or high disease activity). **4)** Baseline comorbid conditions. **5)** Treatment for rheumatic disease: a) glucocorticoids (doses intervals: 0mg, 0-10mg, >10mg), b) nonsteroidal anti-inflammatory drugs [NSAIDs], c) conventional synthetic disease-modifying antirheumatic drugs (csDMARDs); d) targeted synthetic/biologic DMARDs (ts/bDMARDs).

For the identified hospital admission due to COVID-19, physicians collected clinical, laboratory, and treatment data until discharge to describe the evolution of the patients.

Patient characteristics were expressed as mean and standard deviation or median and interquartile range for continuous variables; categorical variables were expressed as percentages. Statistical tests were performed to compare characteristics between patients with COVID-19 asymptomatic and those with symptoms, including hospital admission needed. Continuous variables were analyzed using the Mann-Whitney test, and discrete variables were analyzed using Fisher exact test. Univariable logistic regression analyses were performed to assess differences between asymptomatic or symptomatic cases and also hospital admissions related to COVID-19 risk and covariates. Multivariable logistic regression models (adjusted for age, sex) were run in a stepwise manner to examine the possible effect of sociodemographic, clinical, and therapeutic factors on symptomatic COVID-19 and hospital admissions related to COVID-19 The model also included all other variables with a p<0.1 from the univariable regression analysis. The results were expressed as the odds ratio (OR) with its respective 95% confidence interval.

Study data were collected and managed using REDCap electronic data capture tools hosted at SORCOM.((15)(16)).

The study was approved by the Hospital Ramon y Cajal Institutional Ethics Committee (approval number 136-20).

All analyses were performed in Stata v.13 statistical software (Stata Corp., College Station, TX, USA). A two-tailed *p* value < 0.05 was considered to indicate statistical significance.

## Results

A total of 77 pediatric patients with rheumatic diseases and Covid-19 were included in the study. A 66.23% were diagnosed with a positive SARS-CoV-2 polymerase chain reaction (PCR) test, another 27.27% with a detection of antibodies against SARS-CoV-2 test, and 6.49% had a clinical diagnosis. Fifty-five (71.43%) patients were female (22 male, 28.57 %) and the mean age of patients were 11.88 (SD 4.04) years. Mean time since diagnosis was 5.78 (4.21) years, being the primary diagnosis of patients described in table 1. Most of the patients had a diagnosis of oligoarticular or polyarticular JIA with or without associate uveitis (n=42, 54.54%). Other diagnosis were systemic lupus erythematosus (n=8, 10.38%), systemic JIA (n=5, 6.49%) and monogenic autoinflammatory syndrome (n=5, 6.49%).

**Table 1.** Baseline sociodemographic and clinical characteristics of pediatric patients with Covid-19 and rheumatic diseases.

| Variable | RMD COVID-19 patients<br>n =77 | RMD COVID-19 symptomatic patients<br>n =47 | RMD COVID-19 asymptomatic patients<br>n=30 | p value* |
| --- | --- | --- | --- | --- |
| **Women, n (%)** | 55 (71.43) | 31 (65.95) | 24 (80) | 0.20 |
| **Age, mean (SD), years** | 11.88 (4.04) | 12.04(4.22) | 11.63(3.81) | 0.53 |
| **Clinical remission, n (%)** | 52 (67.53) | 28 (59.57) | 24 (80) | 0.37 |
| **Comorbidities** | 8(10.39) | 8(17.02) | 0 | 0.02 |
| **Diagnosis, n (%)** |  |  |  | 0.53 |
| **Chronic inflammatory arthritis** |  |  |  |  |
| Oligoarticular JIA | 21 (27.27) | 9 (19.14) | 12 (40) |  |
| Oligoarticular JIA with associated uveitis | 9 (11.69) | 5 (10.63) | 4 (13.33) |  |
| Polyarticular JIA | 12 (15.58) | 8 (17.02) | 4 (13.33) |  |
| Enthesitis-related arthritis | 4 (5.19) | 3 (6.38) | 1 (3.33) |  |
| Idiopathic uveitis | 2 (2.60) | 2 (4.25) | 0 |  |
| **Multisystemic autoimmune disorders** |  |  |  |  |
| Systemic lupus erythematosus | 8 (10.38) | 6 (12.76) | 2(6.66) |  |
| Scleroderma | 1 (1.30) | 1 (2.12) | 0 |  |
| Idiopathic inflammatory myopathy | 3(3.90) | 2(4.25) | 1(3.33) |  |
| Pediatric vasculitis | 2 (2.60) | 1 (2.12) | 1 (3.33) |  |
| **Autoinflammatory diseases** |  |  |  |  |
| Systemic JIA | 5(6.49) | 4(8.51) | 1(3.33) |  |
| Chronic recurrent multifocal osteomyelitis | 3(3.90) | 2(4.25) | 1(3.33) |  |
| PFAPA syndrome | 2 (2.60) | 2 (4.25) | 0 |  |
| Monogenic autoinflammatory syndrome | 5(6.49) | 2(4.25) | 3(10) |  |
| **Rheumatic disease treatment, n (%)** |  |  |  |  |
| NSAIDs, n (%) | 3 (3.90) | 2 (4.25) | 1 (3.33) | 0.99 |
| Glucocorticoids, n (%) | 9 (11.69) | 7 (14.89) | 2 (6.66) | 0.46 |
| csDMARDs, n (%) | 35 (45.45) | 22 (46.80) | 13 (43.33) | 0.81 |
| Ts/bDMARDs, n (%) | 40 (51.95) | 24 (51.05) | 16 (53.33) | 0.99 |

\* p<0.05

Few patients had also additional comorbidities: obesity (2 patients), chronic renal disease (2), asthma (1), chronic heart disease (1), previous macrophage activation syndrome (1) and a duplicated collecting system (1) (Table 1).

While 8 of the patients (10.53%) had high RMD activity at the time of COVID-19 diagnosis, most (89.47%) had low activity or were in clinical remission.

Prior to COVID-19 infection, only nine of the patients were taking glucocorticoids (11.68%) and 3 were taking NSAIDs on a regular basis (3.90%). Almost half of patients were taking csDMARDs, mainly methotrexate (N=26, 33.77%) and antimalarials (N=7, 9.09%) and mycophenolate mofetil (N=6, 7.79%). 51.95% were taking ts/bDMARDs, mainly anti-TNF therapy, being etanercept the most frequently prescribed (19.48%), followed by adalimumab (14.29%). Only one patient was taking a Janus-Kinasa (JAK) inhibitor. Additionally, 3 patients were taking Angiotensin-converting-enzyme (ACE) inhibitors and two were anticoagulated.

Of the 77 patients, 47 had at least one COVID-19-related symptom (mainly fever, headache, cough), while 30 patients were asymptomatic. Symptomatic patients were older (14 [8.5-16] vs 12 [9.5-15]) than asymptomatic ones, without significant differences. None of the patients was diagnosed with pediatric multisystemic inflammatory syndrome. 81.52% of all patients maintained all their immunosuppressive treatment without changes. A 19.48% of patients went to the emergency room due to their symptoms.

Multisystemic autoimmune disorders had a higher percentage of symptomatic patients (71.4%), followed by autoinflammatory diseases, with 66.6%. However, there were no significant differences between diseases groups.

As we show in table 2, most patients had mild disease, with fever as the most prevalent symptom. None critical disease was found. Hospital admission was required in 6 patients (7.79% of all patients), with a median of 5 days (IQR 2-20) and pediatric intensive care unit admission in one. In each RMD diagnostic category there were two cases of hospital admission,. Half of the admitted patients were on bDMARDS (2 Infliximab, 1 Canakinumab), being their sociodemographic and clinical characteristics described in Table 3.

**Table 2.** Characteristics of patients with symptomatic COVID-19 related symptoms.

| **Variable** | **RMD Covid-19 patients**<br>**n =47** |
| --- | --- |
| **Women, n (%)** | 31 (65.95) |
| **Age, mean (SD), years** | 12.04(4.22) |
| **Activity of the RMD** |  |
| Mild | 41 (87.23) |
| Moderate | 5 (10.63) |
| Severe | 1 (2.12) |
| **COVID-19 symptoms** |  |
| Dyspnea | 2 (4.25) |
| Fatigue | 10 (21.27) |
| Chest pain | 5 (10.63) |
| Cough | 17 (36.17) |
| Abdominal pain | 2 (4.25) |
| Diarrhoea/vomiting | 10 (21.27) |
| Fever | 45 (95.74) |
| Anosmia | 10 (21.27) |
| Dysgeusia | 12 (25.53) |
| Joint pain | 8 (17.02) |
| Headache | 20 (42.55) |
| Odynophagia | 16 (34.04) |
| **Hospital admission required, n (%)** | 6 (12.76) |

**Table 3.** Characteristics of the admitted patients with COVID-19.

| Variable | COVID-19<br>admitted patients<br>n =6 |
| --- | --- |
| **Women, n (%)** | 5 (83.33) |
| **Age, median (IQR), years** | 14.5 (4-16) |
| **Diagnosis, n (%)** |  |
| Systemic JIA | 1 (16.6) |
| Oligoarticular JIA with associated uveitis | 2 (28.8) |
| Systemic lupus erythematosus | 1 (16.6) |
| Vasculitis | 1 (16.6) |
| Autoinflammatory syndrome | 1 (16.6) |
| **COVID-19–related treatments during admission, n (%)** |  |
| Hydroxychloroquine | 1 (16.6) |
| Glucocorticoids | 2 (28.8) |
| Azithromycin | 1 (16.6) |
| Lopinavir/ritonavir | 1 (16.6) |
| **Admission duration, median (IQR), (days)** | 5 (2-15) |

The only patient who required admission to a Pediatric Intensive Care Unit had a recent diagnosis of systemic JIA in treatment with Canakinumab and glucocorticoids (> 10 mg prednisone/day), and had associated comorbidities. This patient developed pneumonia and pleural effusion and was treated with antibiotic therapy and lopinavir/ritonavir. A catheter-associated right vein iliac thrombosis appeared as a probably multifactorial complication. A few days later the patient’s condition improved, and the recovery was completely at discharge.

No laboratory or imaging tests were performed for asymptomatic and mild symptomatic patients. All of them were followed up without specific treatment.

The multivariable logistic regression model for symptomatic infection was adjusted for age, sex and previous comorbidities. None of variables included in the final model reached statistical significance. Comorbidities dropped from the final model because predict failure perfectly. Exposure to DMARDs [biological (OR: 1.02 [0.34-3.09]; p=0.95) or synthetic (OR: 1.24 [0.44-3.46]; p=0.67)]), and exposure to glucocorticoids (OR: 1.27 [0.47-3.47]; p=0.62)), did not achieve statistical significance and finally dropped from the last model. Although, long-term treatment with antimalarials seemed to be protector, it did not reach significance in the final model (OR: 0.11 [0.007-1.88]; p=0.13). (Table 4).

**Table 4.** Multivariable logistic regression models. Associated factors for symptomatic infection and hospital admission.

|  | Symptomatic<br>infection<br>Odd ratio (95% CI) | Hospital Admission<br>Odd ratio (95% CI) |
| --- | --- | --- |
| **Age** | 1.04 (0.91 to 1.20) | 1.06 (0.78 to 1.46) |
| **Sex** | 2.03 (0.65 to 6.34) | 0.38 (0.03 to 4.15) |
| **Diagnosis: Multisystemic autoimmune disorders vs chronic inflammatory arthritis** | 1.55 (0.38 to 6.27) | 2.09 (0.19 to 22.01) |
| **Diagnosis: Autoinflammatory diseases vs chronic inflammatory arthritis** | 1.42 (0.33 to 6.09) | 3.30 (0.16 to 66.15) |
| **Glucocorticoids** | 1.27 (0.47 to 3.47) | 3.51 (1.04 to 11.84)* |

\* p<0.05

In the multivariable logistic regression adjusted model for hospital admission, exposure to more doses of glucocorticoids was the only variable associated with higher odds of hospital admission. Again, the exposure to DMARDs, -regardless of whether they were biological (OR: 1.97 [0.23-16.57]; p=0.53) or synthetic (OR: 1.20 [0.15-9.56]; p=0.86)-, did not reach statistical significance in the final model. Comorbidities also dropped from the final model (OR: 3.45 [0.41-28.48]; p=0.25). (Table 4).

## Discussion

Our study aims to shed light on pediatric rheumatologists’ concerns regarding their patients. This study conducted in six tertiary care hospital presents the baseline characteristics and outcomes of pediatric patients with rheumatic diseases who were infected with SARS-CoV-2.

We found that, in the COVID-SORCOM register, a 61% of pediatric patients with RMD and COVID-19 have a symptomatic infection. In concordance with our observations, fever, headache and cough were the predominant clinical features at presentation, with similar rates observed in a multinational, multicenter study on pediatric COVID-19 (17).

Presence of a pre-existing medical comorbidities, was associated with increased likelihood of having a symptomatic disease in patients with RMD.

We also describe that, less than 8% of the patients required hospital admission. Our data also reflect the uncertainties regarding drug treatment options for COVID-19 with different protocols as knowledge of the disease progressed. In general, admitted patients received few experimental treatments for COVID-19 and in almost all cases the symptoms remitted without serious or prolonged complications. Our data shows that patients exposed to disease-modifying agents do not seem to be at higher risk of severity or hospital admission related to COVID-19. Nevertheless, our results suggest that pediatric patients with RMD and chronic prior exposure to glucocorticoids and even to more doses had more probability of hospital admissions, regardless of other factors. Although we have to consider the limited number of patients in our study, our results are in concordance with data reported elsewhere in adults patients with RMD and COVID-19 (6)(11). The data about glucocorticoid use are also complex and challenging to interpret, taking into account that pediatric rheumatologist always try to maintain the lowest possible dose and that those with higher doses usually have a more serious disease or higher diseases activity.

The possibility of a worse prognosis in patients with rituximab has recently been discussed (18)(19)(20). In our cohort there was only a patient with systemic lupus erythematosus that has received rituximab in the previous 3 months of infection and, although had symptomatic disease, did not required hospital admission. This patient also were using hydroxychloroquine and mycophenolate mofetil, being the latter suspended until a complete recovery was achieved.

Although available data support that children with RMD undergoing immunomodulatory treatment do not have a severe COVID-19, conventional clinical practice is the temporary interruption of immunomodulatory treatment when there is a concomitant infection (21). In our cohort, in asymptomatic infections, most of the physicians maintained the immunomodulatory treatment. Also in most of the patient with symptomatic COVID-19, DMARDs (biological or synthetic) were maintained. There is no evidence that continuing DMARDs during COVID-19 can alter the clinical course of SARS-CoV-2 infection and the benefits of keeping the disease inactive outweighed the risks (18)(21).

The type of diagnosis seems to play an important role in the probability of hospital admission in adult patients with RMD and COVID-19, and patients with systemic autoimmune conditions seem to have the highest risk compared with chronic inflammatory arthritis(6)(18). However, we have not been able to replicate these results in our pediatric population and we did not find a higher risk for any of the outcomes.

The rest of the admitted patients did not develop associated complications, except for an adolescent girl with systemic lupus erythematosus and secondary antiphospholipid syndrome who had an episode of a deep vein thrombosis in a lower limb. Thrombotic complications seem very uncommon in children with SARS-CoV-2, however in adolescents with previous thrombotic risk factors thromboprophylaxis must be considered (22).

A large majority of children with asymptomatic disease were in remission. This might reflects the importance of keeping the disease under control to avoid symptomatic COVID-19. Patients with high or moderate disease activity may be at risk for superimposed infections including SARS-CoV-2.

Strengths of our study include a large analysis of pediatric patients with RMD and COVID-19 during the peak pandemic. All case data were entered by their own rheumatology healthcare providers. The register includes cases from the main pediatric rheumatic units in the Community of Madrid suggesting that our findings are more generalizable than single-centre studies. The registry collects information on specific RMD diagnoses, which to date have not been captured in other case series of COVID-19.

Despite these strengths, there are important limitations to these registry data. The COVID-SORCOM registry is voluntary and does not capture all cases of COVID-19 in pediatric patients with RMD. This approach to data collection places limitations on causal conclusions and temporal relationships and therefore we can only make limited inferences based on our results. Due to the database design and inherent reporting bias the data cannot be used to comment on the incidence of COVID-19 in this patient population.

The limited size of the sample makes it a descriptive study with exploratory analyzes, but still gives us detailed information about this population, and brings us closer to preliminary results that are in line with what has been published in the adult population with RMD, helping to make decision-making and patient monitoring. Although further studies are required, children with RMD or receiving immunomodulatory therapies don’t seem to have a higher risk for severe COVID-19 and preventive measures are similar to those counseled to the general population(23)(21).

In conclusion, overall outcomes in children and adolescents with RMD appear to be generally good, with mild infection. However, we should keep in mind that children are not immune to COVID-19, and although rarely, the disease could be severe and complicated. Comorbidities and the use of glucocorticoids could be considered as risk factors in the RMD pediatric population as well as in adults. These factors could have a combined overall effect, so special attention should be paid to patients with associated comorbidities and glucocorticoids treatment.

## Conclusions

Overall, our data found no differences in COVID-19 outcomes between children-onset rheumatic diseases; however, some factors may increase the likelihood that patients with symptomatic infection may develop a complication or require hospital admission. In this real-world clinical registry, our results are in concordance with data reported elsewhere in adult patients with RMD and COVID-19, warning to take precautions with patients with thrombotic risk, with comorbidities or with previous exposure to corticosteroids. This study provides novel results in children-onset rheumatic disease patients regarding susceptibility to moderate-severe infection related to COVID-19.

## Data Availability

The datasets generated and analyzed for the present study are available from the corresponding author on reasonable request.

## Abbreviations

ACE: angiotensin-converting-enzyme inhibitors
CAPS: cryopyrin-associated periodic syndrome
COVID-19: Coronavirus disease 2019
DMARD: disease-modifying anti-rheumatic drug cs
DMARDs: classic DMARDs
ts/bDMARDs: targeted synthetic or biologic DMARDs
FMF: familial Mediterranean fever
JAK: Janus-Kinasa
JIA: juvenile idiopathic arthritis
HIDS: hyperimmunoglobulin-D syndrome
IQR: interquartile Range
MIS-C: multisystem inflammatory syndrome in children
MKD: mevalonate kinase deficiency
NSAID: nonsteroidal anti-inflammatory drugs
PCR: polymerase chain reaction
PFAPA: periodic fever, adenopathy, pharyngitis and aphthous stomatitis
RMD: rheumatic and musculoskeletal diseases
SARS: severe acute respiratory syndrome
TNF: tumor necrosis factor
TRAPS: TNF- receptor associated periodic syndrome

## Acknowledgements

We wish to thank all rheumatology providers who entered data into the registry.

## Author information

### Contributions

DC and LL wrote the manuscript, which was critically revised by AB, JB and LA. All authors contributed to manuscript revisions, and read and approved the submitted version.

## Ethics declarations

### Consent for publication

N/A

